# Modifying reusable elastomeric respirators to utilise breathing system filters with 3D printed adapters, a safe alternative to N95 during COVID-19

**DOI:** 10.1101/2020.04.10.20061291

**Authors:** D.C.Y. Liu, T.H. Koo, J.K.K. Wong, Y.H. Wong, K.S.C. Fung, Y. Chan, H.S. Lim

## Abstract

The COVID-19 pandemic has caused a worldwide shortage of personal protective equipment including N95 and FFP3 respirators. Reusable elastomeric respirators are suitable alternatives when used with compatible filters. These filters may be difficult to source and elastomeric respirators are not recommended for surgical use as the exhaled air is not filtered. Breathing system filters are routinely used in anaesthetic circuits to filter virus and bacteria. In this study, we designed 3D printed adapters that allowed elastomeric respirators to utilise breathing system filters and made simple modifications to the respirators to filter exhaled breaths. We then evaluated the performance and safety of our modified elastomeric respirators with quantitative fit tests.

We recruited 8 volunteers to perform quantitative fit tests. Fit factors, respiratory rate and end-tidal carbon dioxide were recorded before and after wearing the modified respirators for 1 hour. All 8 volunteers obtained fit factors of 200+, the maximum achievable, for all tests exercises in all fit tests. The mean (range) end-tidal carbon dioxide was 4.5 (3.9-5.5) kPa and 4.6 (range 4.1-5.3) kPa before and after 1 hour of usage. The mean (range) respiratory rate was 16.5 (11-24) min^-1^ and 17.4 (15-22) min^-1^ before and after 1 hour of usage. Four (50%) did not experience any subjective discomfort while 2 (25%) reported pressure on the face, 1 (12.5%) reported exhalation resistance and 1 (12.5%) reported transient dizziness with exertion. Breathing system filters combined with properly fitted reusable elastomeric respirators is a safe alternative to N95 during the COVID-19 pandemic.

## Introduction

The COVID-19 pandemic has caused a worldwide surge in the usage of personal protective equipment including N95 and FFP3 respirators. These respirators are recommended when performing aerosol generating procedures including intubation and extubation[1,2]. For anaesthetists who perform aerosol generating procedures regularly, the critical shortage of these masks is a pressing issue. Reusable elastomeric respirators are suitable alternatives when used with compatible National Institute for Occupational Safety and Health (NIOSH) or CE approved filters such as P100 and P3[1,3]. It has been shown that healthcare personnel can be rapidly fit tested and trained to use them[4]. These filters may also be short in supply however, and elastomeric respirators are not recommended for surgical use as the exhaled air is not filtered[1], limiting their use in the operating theatre.

Breathing system filters are routinely used in anaesthetic circuits and ventilators. They are designed to filter virus and bacteria[5], and are readily available in any operating theatre or intensive care unit. In this study, we designed 3D printed adapters that allowed 3M elastomeric respirators (St Paul, Minnesota, USA) to utilise breathing system filters and made simple modifications to the elastomeric respirators to filter exhaled breaths. We then evaluated the performance and safety of our modified elastomeric respirators with quantitative fit tests.

## Methods

We recruited 8 volunteers after ethics approval by the Hospital Authority Research Ethics Committee and written informed consent. An adapter and a cap were designed with SolidWorks (Dassault Systèmes, Vélizy-Villacoublay, France) and printed with an Ultimaker S5 3D printer (Utrecht, Netherlands) using polylactic acid (Premium PLA, Formfutura BV, Nijmegen, Gelderland, Netherlands). The adapter was designed to fit the 3M Bayonet connector and to accommodate a standard 22mm outer diameter connector (figure 1). The cap was designed to seal off the 3M Bayonet connector. The 3D printed adapter was attached to 3M 6200 (medium) and 7501 (small) respirators and modified into configuration 1 (inhalation through breathing system filter, exhalation through exhalation valve) and configuration 2 (inhalation and exhalation through breathing system filter). For configuration 2, the inhalation valves were removed (figure 2) and the exhalation valve was sealed by wedging a piece of plastic (5×7cm on 3M 7501, 5×5cm on 3M 6200) cut out from a zipper storage bag between the exhalation valve and the head harness assembly (figure 3-4). Undis BVF-02 (Shaoxing, Zhejiang, China) BSFs were used for testing.

**Figure 1.**
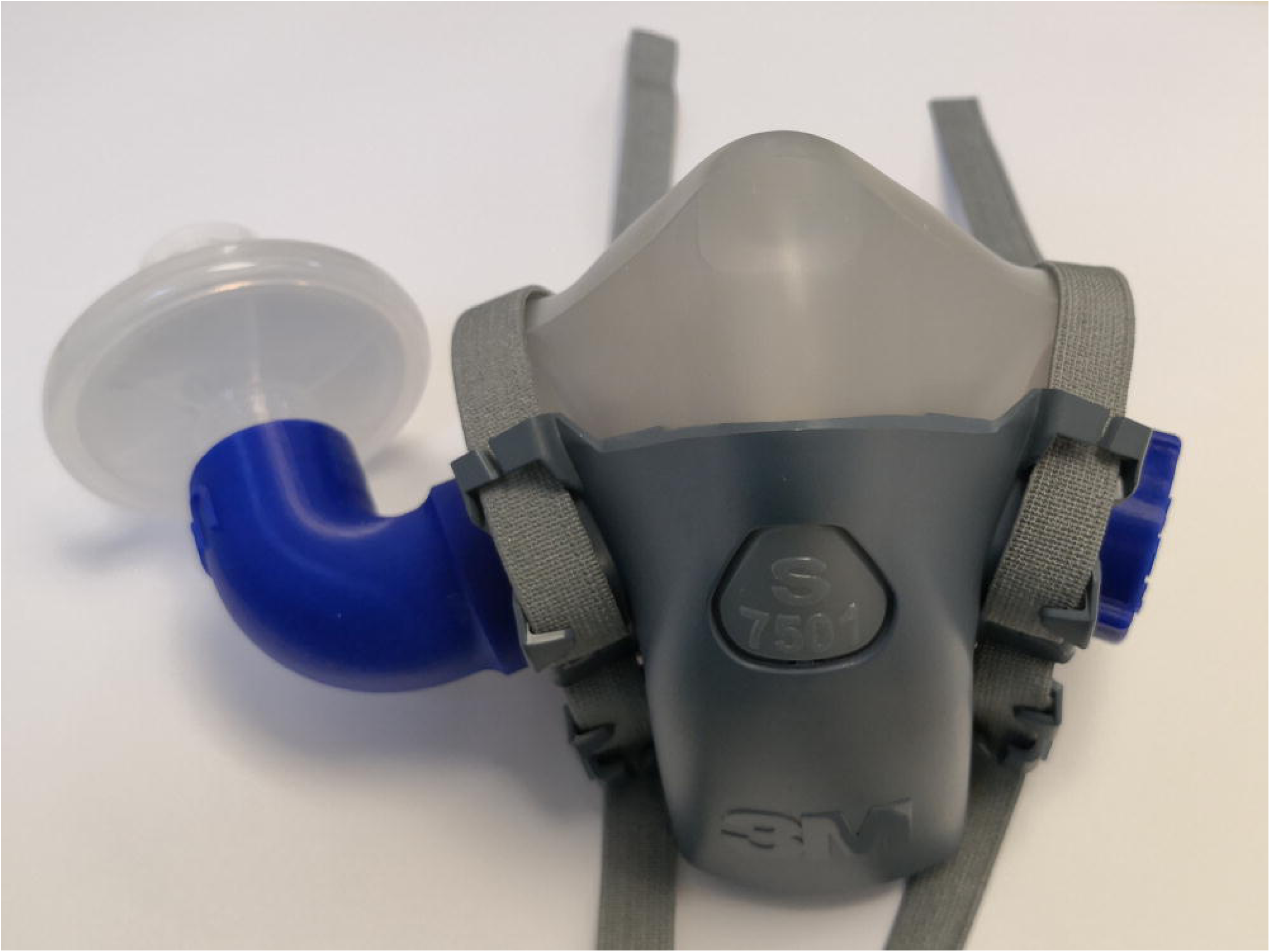
3M 7501 with 3D printed adapter, cap and breathing system filter installed

**Figure 2.**
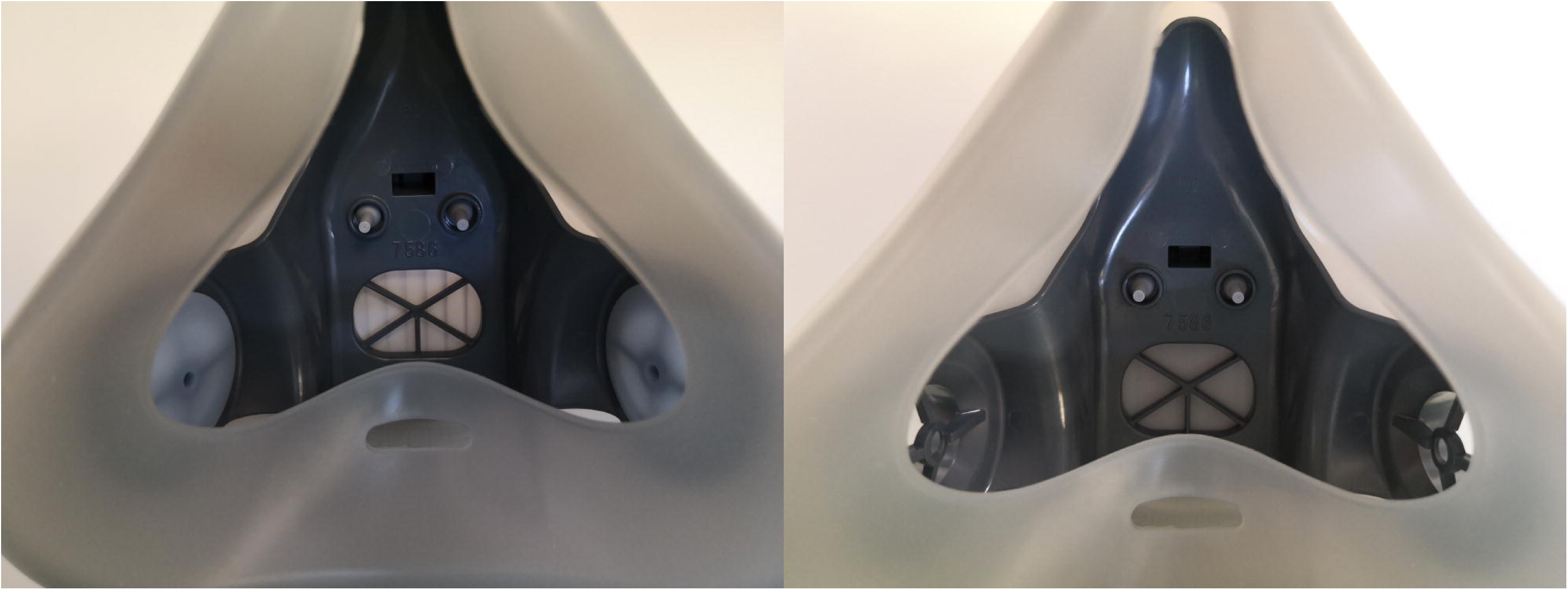
3M 7501 with inhalation valves intact (left) and removed (right)

**Figure 3.**
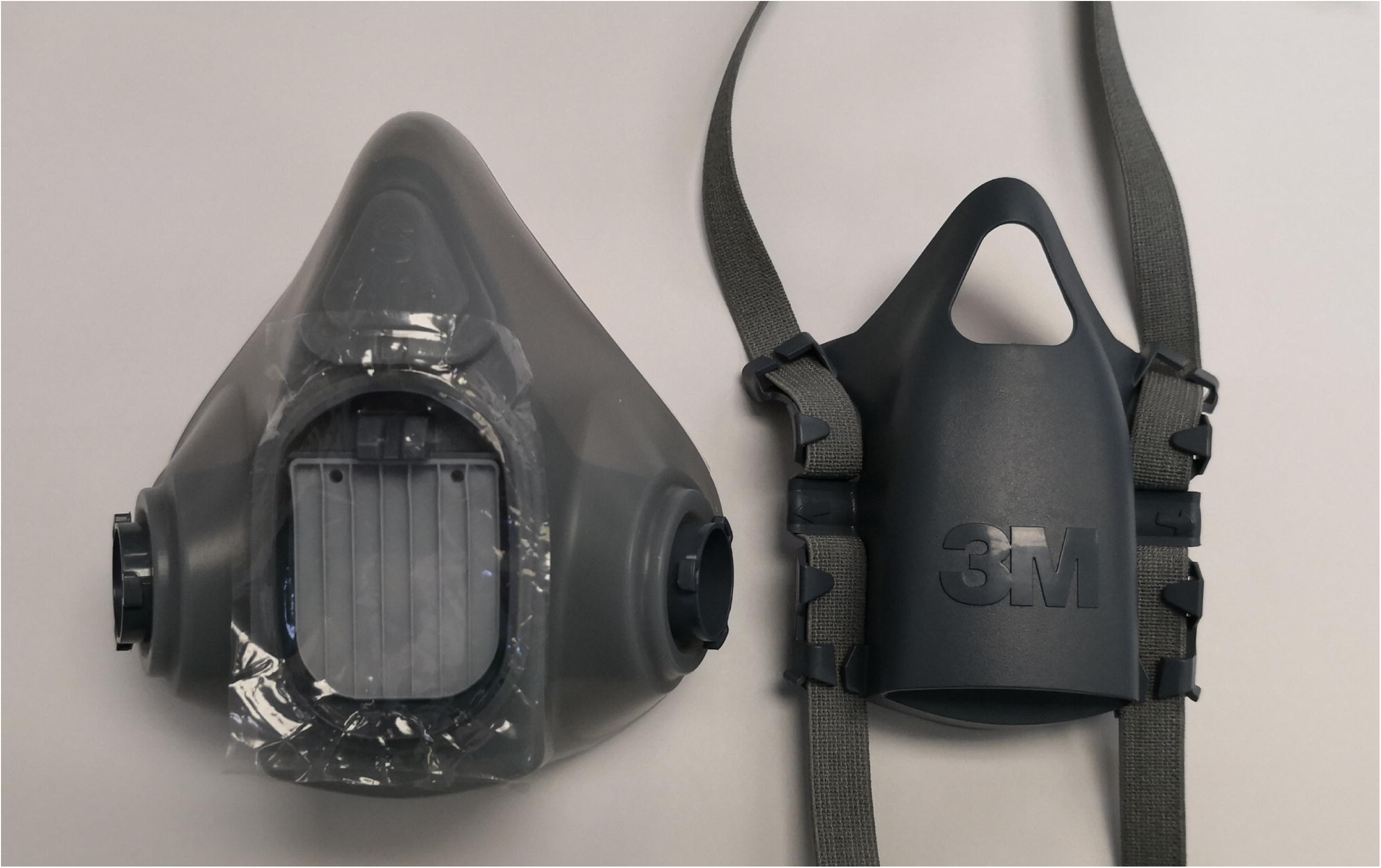
3M 7501 with a piece of 5×7cm plastic, head harness assembly detached

**Figure 4.**
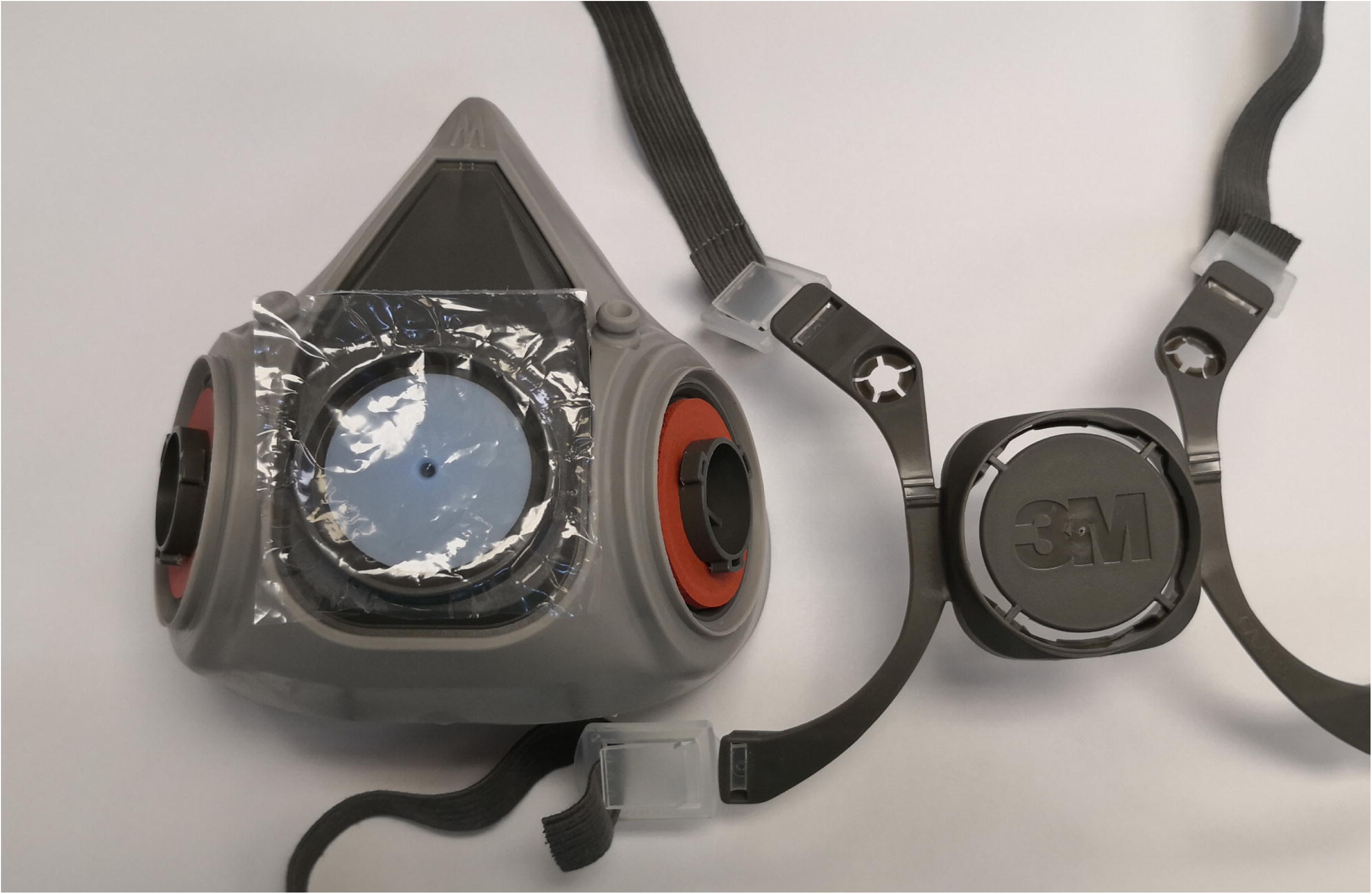
3M 6200 with a piece of 5×5cm plastic, head harness assembly detached

The negative pressure seal check was performed by inhaling while the breathing system filter was covered with the volunteer’s hand. The positive pressure seal check was performed by exhaling while the exhalation opening (configuration 1) or breathing system filter (configuration 2) was covered with the volunteer’s hand. Leak was detected subjectively by the volunteers. Real time fit factor test was performed on a PortaCount Pro+ 8038 fit tester (TSI, Shoreview, Minnesota, USA) via a 3M 601 fit test adapter installed between the elastomeric respirator and the 3D printed cap. These initial tests were determined as satisfactory if no leaks were detected during pressure seal checks and stable fit factors were obtained during the real time fit factor tests. If the initial tests were unsatisfactory, the volunteers were fitted with alternate sized respirators. Following satisfactory initial tests, the volunteers performed full standardised quantitative fit testing[6] using the manufacturer’s N95-Companion protocol[7]. The volunteers performed test exercises consisting of normal breathing, deep breathing, head side to side, head up and down, talking out loud, grimace, bend and touch toes and normal breathing. Fit testing was done 3 times, first in configuration 1, second in configuration 2, and third in configuration 2 after wearing the respirators for 1 hour. End-tidal carbon dioxide and respiratory rate were measured with a Capnocheck II carbon dioxide detector (Smiths Medical, Minneapolis, Minnesota, USA) before and after wearing the respirators for 1 hour.

## Results

Eight volunteers (5 male, 3 female) completed the study. The mean (range) age was 44.1 (34-58) years. All 8 passed the pressure seal checks. Four males failed the real time fit factor test wearing medium respirators with unstable fit factor during head movement. They were subsequently fitted with small respirators and passed the initial tests. All 3 females passed the initial tests with small respirators.

All 8 volunteers obtained fit factors of 200+, the maximum achievable, for all tests exercises in all 3 fit tests. The mean (range) end-tidal carbon dioxide was 4.5 (3.9-5.5) kPa and 4.6 (range 4.1-5.3) kPa before and after 1 hour of usage. The mean (range) respiratory rate was 16.5 (11-24) min^-1^ and 17.4 (15-22) min^-1^ before and after 1 hour of usage. Four (50%) did not experience any subjective discomfort while 2 (25%) reported pressure on the face, 1 (12.5%) reported exhalation resistance and 1 (12.5%) reported transient dizziness with exertion.

## Discussion

This study demonstrated that utilising breathing system filters on modified reusable elasomeric respirators is safe and feasible. Excellent quantitative fit test results were demonstrated and this persisted following 1 hour of usage. Exhalation through breathing system filters was well tolerated and carbon dioxide rebreathing due to equipment dead space was not of concern as evidenced by the normal end-tidal carbon dioxide.

3M elastomeric respirators are designed to undergo repeated disinfection. Our study showed that the polylactic acid material used for 3D printing is airtight when subjected to pressures encountered during normal breathing, as no leak was detected. Bleach and alcohol can be used to disinfect the 3D printed adapters according to the manufacturer. Further studies are required in order to evaluate their resistance to repeated disinfection and their physical durability with usage. Other materials may be used for 3D printing as long as they are airtight, non-toxic and resistant to disinfection. We designed a 90-degree angle in our adapter in order to facilitate the use of face shields and to reduce the risk of contamination from splashing. The cost to print an adapter and a cap is approximately £3. We designed another version that requires less material and time to print and has less equipment dead space (figure 5). This version obstructs the use of face shields slightly and was not tested in the study. This version with a cap cost around £1.8 to print.

**Figure 5.**
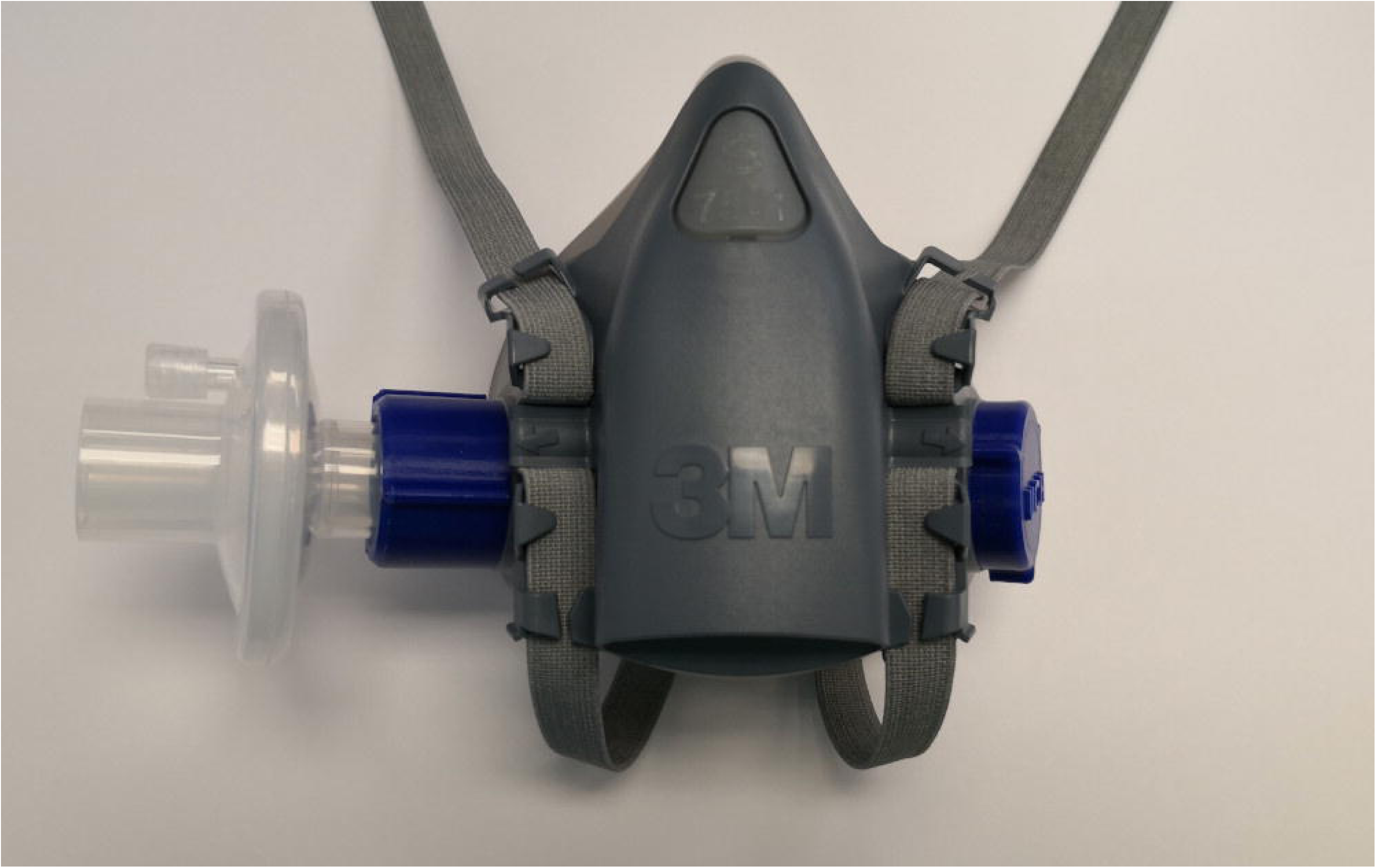
3M 7501 with an alternate 3D printed adapter, cap and breathing system filter installed

Although the elastomeric respirators were well tolerated, the slight increase in air resistance during exhalation through the filters is noticeable. This can be overcome by using 2 filters and prolonged use may be feasible. It is worth noting that during general anaesthesia, which produces respiratory depression, patients regularly breathe spontaneously through a single filter and encounter similar resistance for several hours without much detrimental physiological consequences. Wearing these well-fitted, tightly sealed respirators will expectedly affect communication, especially when the exhalation valve is sealed. We have found that although the voice is mumbled and lower in volume, it is possible to understand the user’s speech in a quiet environment.

There are two types of breathing system filters, electrostatic and mechanical (also known as pleated hydrophobic). Either one may be combined with a heat and moisture exchanger. Advantages of electrostatic filters include low resistance, light weight, small size and low cost, while disadvantages include vulnerability to liquid (and any microbe contained in the liquid) and lower filtration efficiency[5]. Mechanical filters on the other hand, have higher filtration efficiency than electrostatic filters in general and are more resistant to liquid. Disadvantages include higher resistance, heavier weight, larger size and higher cost[8]. The Undis BVF-02 filter used in our study is of the electrostatic type and does not have a heat and moisture exchanger included, it has a viral and bacterial filtration efficiency of >99.9999%[9]. We chose this filter as it has low resistance and a heat and moisture exchanger is not required.

A fit factor of >100 constitutes a pass when fit testing half facepiece respirators[6]. During preliminary testing on the authors, we found that despite obtaining perfect fit factors of 200+ with the N95-Companion protocol, the fit factors become close to 0 when the P100 protocol is used. This phenomenon remained the same when we tested another model of electrostatic heat and moisture exchanger filter, Medtronic DAR 352U5877 (Minneapolis, Minnesota, USA). We then tested two models of mechanical filters, Medtronic DAR 351U5410 and Pall Ultipor 100 (Port Washington, New York, USA), both of which passed the N95-Companion and P100 protocols. This is explained by how the PortaCount Pro+ 8038 fit tester detects a leak in the N95-Companion protocol, which is valid for measuring the fit of any respirator using filter media that is similar to NIOSH approved N95 respirators[10]. Electrostatic attraction is a very important mechanism for the filtration efficiency of polypropylene used in NIOSH approved N95 respirators[11,12], this adds support to the validity of the N95-Companion protocol to detect leaks when using electrostatic breathing system filters.

All of the filters tested had viral and bacterial filtration efficiencies of >99.999% according to the manufacturers[9,13–15]. Different manufacturers may use different methods to calculate these numbers however, and the results of the standardised sodium chloride particles penetration test[16] are often not reported. Electrostatic filters may have high viral and bacterial filtration efficiency but they do not filter >99% of particles measured by the fit tester, hence failing the P100 protocol. For maximum level of protection, mechanical filters with the highest sodium chloride particles filtration efficiency can be considered. Nonetheless, quantitative fit testing using the N95-Companion protocol is currently the gold standard used to detect leaks from N95 respirators. Quantitative fit test should be mandatory before using reusable elastomeric respirators as a significant proportion of volunteers failed the real time fit factor test despite having satisfactory pressure seal checks, this is in line with published guidelines[3,6].

For easy repeatability during our study, we sealed the exhalation opening with a piece of plastic. This may not be a very reliable method for clinical usage. Should the piece of plastic fail however, the safety of the user will not compromised as the exhalation valve will still be intact and the inhaled air remains filtered. We are currently experimenting with a different method of sealing the exhalation opening by wedging medical grade silicone and a 3D printed spacer between the exhalation opening and head harness assembly. If the supply of surgical masks is not of concern and use in a sterile field is required, an alternate method would be to leave the valves unmodified and to wear a surgical mask on top of the elastomeric respirator to filter exhaled breaths.

Limitations of our study include our small sample size, short duration of respirator usage and that only 1 model of filter was tested. We chose to test the respirators for 1 hour as most aerosol generating procedures can be completed within this timeframe, after which a lower level of respiratory protection with surgical masks may be adequate. Preliminary testing on one of the authors suggests that the respirator can be worn for 4 hours without significant change in respiratory rate and end-tidal carbon dioxide but further studies are required to evaluate their safety and effectiveness after prolonged usage. Most manufacturers recommend that their breathing system filters can be used for up to 24 hours[8].

In conclusion, we successfully converted a piece of anaesthetic equipment into personal protective equipment that can be used in sterile fields. When N95/FFP3 respirators and NIOSH/CE approved filters are difficult to source, breathing system filters combined with properly fitted reusable elastomeric respirators is a safe alternative.

## Data Availability

The datasets generated during and/or analysed during the current study are available from the corresponding author on reasonable request.

